# Small RNA signatures of the anterior cruciate ligament from patients with knee joint osteoarthritis

**DOI:** 10.1101/2020.05.14.20101048

**Authors:** Yalda A. Kharaz, Yongxiang Fang, Tim Welting, Mandy Peffers, Eithne J. Comerford

## Abstract

The anterior cruciate ligaments are susceptible to degeneration, resulting in pain, reduced mobility and development of the degenerative joint disease osteoarthritis. There is currently a paucity of knowledge on how anterior cruciate ligament degeneration and disease can lead to osteoarthritis. Small non-coding RNAs (sncRNAs), such as microRNAs, and small nucleolar RNA, are important regulators of gene expression. We aimed to identify sncRNA profiles of human anterior cruciate ligaments to provide novel insights into their roles in osteoarthritis.

RNA was extracted from the anterior cruciate ligaments of non-osteoarthritic knee joints (control) and end-stage osteoarthritis knee joints, used for small RNA sequencing and significantly differentially expressed sncRNAs defined. Bioinformatic analysis was undertaken on the differentially expressed miRNAs and their putative target mRNAs to investigate pathways and biological processes affected.

Our analysis identified 184 sncRNA that were differentially expressed between control ACLs derived from osteoarthritic joints with a false discovery adjusted p value<0.05; 68 small nucleolar RNAs, 26 small nuclear RNAs and 90 microRNAs. We identified both novel and previously identified (miR-206, –101, –365 and –29b and –29c) osteoarthritis-related microRNAs and other sncRNAs (including SNORD74, SNORD114, SNORD72) differentially expressed in ligaments derived from osteoarthritic joints. Significant cellular functions deduced by the differentially small nuclear RNAs and 90 microRNAs. We identified expressed miRNAs included differentiation of muscle (P<0.001), inflammation (P<1.42E-10), proliferation of chondrocytes (P<0.03), fibrosis (P<0.001) and cell viability (P<0.03). Putative mRNAs were associated with the canonical pathways ‘Hepatic Fibrosis Signalling’ (P<3.7E-32), and ‘Osteoarthritis’ (P<2.2E-23). Biological processes included apoptosis (P<1.7E-85), fibrosis (P<1.2E-79), inflammation (P<3.4E-88), necrosis (P<7.2E-88) and angiogenesis (P<5.7E-101).

SncRNAs are important regulators of anterior cruciate disease during osteoarthritis and may be used as therapeutic targets to prevent and manage anterior cruciate ligament disease and the resultant osteoarthritis.

## INTRODUCTION

Ligaments are resilient connective tissues essential for bone-to-bone connections within joints [1]. The anterior cruciate ligament (ACL) is the most commonly damaged ligament [2] with an incidence of approximately 68.6 ACL ruptures per 100,000 people [3] resulting in considerable social and economic costs [4, 5]. In the USA alone, there are approximately 100,000–175,000 ACLs surgeries per year, with cost exceeding of 2 billion dollars [6, 7]. ACL injuries can also lead to significant functional impairment in athletes, muscle atrophy and weakness, joint instability, meniscal lesions, and are associated with development of osteoarthritis (OA) [8, 9]. In the case of the knee joint, more than 50% of ACL injury patients eventually develop OA with the degree and progression of disease being accelerated in these cases [10, 11]. Reports demonstrate that there is an association between ACL degeneration and subsequent knee OA, suggesting the importance of ACL degradation in OA pathogenesis [12].

There is currently substantial interest in the area of epigenetic regulation in ageing, disease, and repair mechanisms in musculoskeletal tissues such as muscle [13, 14], cartilage [15, 16], tendon [17, 18] and ligament [19, 20]. Epigenetics is the study of changes in gene expression that do not derive from changes to the genetic code itself [21]. Insufficient exploration of the epigenetic changes in diseased ACL has been undertaken. One class of epigenetic molecules are small non-coding RNAs (sncRNAs) which include microRNAs (miRNAs or miRs), small nucleolar RNAs (snoRNAs) and small nuclear RNAs (snRNAs) These are functional RNA molecules that are transcribed from DNA but do not translate into proteins and are emerging as important regulators of gene expression before and after protein transcription. Their aberrant expression profiles in musculoskeletal conditions such as ACL injury are expected to be associated with cellular dysfunction and disease development [22]. We have previously identified changes in the sncRNA profiles in ageing and OA human and equine cartilage [16, 23, 24], ageing human and equine tendon [25, 26] and ageing and OA murine joints and serum [27]. Identifying sncRNAs associated with ACL degradation and comprehending their role in OA could have an enormous impact on the understanding of its pathogenesis and future management.

To date, little is known about the sncRNA changes in diseased ACL. We hypothesise that sncRNA expression is altered in ACLs derived from OA joints and that their identification may elucidate the underlying mechanisms of ACL degeneration. This information could then provide potential diagnostic markers and enable future therapeutic targets to treat ACL degeneration, facilitating prompt positive intervention in the associated development of OA.

## MATERIALS AND METHODS

All reagents were from ThermoFisher Scientific, unless otherwise stated.

### Sample collection

ACLs from non-OA, apparently healthy knee joints (control) (n=4) were obtained from a commercial biobank (Articular Engineering). Ethical approval for the purchase human ACL tissue was granted by the Central University Research Ethics Committee C, University of Liverpool (RETH4721). Diseased OA ACLs were obtained from the knee joints of patients undergoing total knee arthroplasty for end-stage OA treatment (n=4). Fully informed patient consent was given for the use of these samples under the institutional ethical approval (Maastricht University Medical Centre approval IDs: MUMC 2017–0183). Samples were collected in RNAlater and stored at –80°C until used.

### RNA extraction

RNA was extracted from ACL tissues once pulverised into a powder with a dismembranator (Mikro-S, Sartorius, Melsungen, Germany) under liquid nitrogen. Total RNA was extracted using the miRNeasy kit (Qiagen, Manchester, UK) according to the manufacturer’s instructions. The RNA samples were quantified using a Nanodrop spectrophotometer (NanoDrop Technologies, Wilmington, USA). The integrity of the RNA was assessed on the Agilent 2100 Bioanalyzer (Agilent, Stockport, UK) using an RNA Pico chip (Agilent, Stockport, UK).

### Small RNA-Sequencing analysis: cDNA library preparation and sequencing

1000ng RNA per sample was submitted for library preparation using NEBNext® Small RNA Library Prep Set for Illumina (New England Biosciences (NEB), Ipswich, USA) but with the addition of a Cap-Clip™ Acid Pyrophosphatase (Cell script, Madison, USA) step to remove any 5’ cap structures on some snoRNAs [27] and size selected using a range 120–300bp (including adapters). This enabled both miRNAs and snoRNAs to be identified using a non-biased approach. The pooled libraries were sequenced on an Illumina HiSeq4000 platform with version 1 chemistry to generate 2 × 150 bp paired-end reads.

### Data processing

Sequence data were processed through a number of steps to obtain non-coding RNA expression values including; basecalling and de-multiplexing of indexed reads using CASAVA version 1.8.2; adapter and quality trimming using Cutadapt version 1.2.1 [28] and Sickle version 1.200 to obtain fastq files of trimmed reads; aligning reads to human genome reference sequences (release 95) from Ensembl using Tophat version 2.0.10 [29] with option “–g 1”; counting aligned reads using HTSeq-count [30] against the features defined in human genome GTF file (release 95). The features whose biotype belonged to the gene categories such as miRNA, snoRNA, and snRNA were extracted.

Differential expression (DE) analysis was performed in R using package DESeq2 [31]. The processes and technical details of the analysis include; assessing data variation and detecting outlier samples through comparing variations of within and between sample groups using principle component analysis (PCA) and correlation analysis; handling library size variation using DESeq2 default method; formulating data variation using negative binomial distributions; modelling data using a generalised linear model; computing log fold change (logFC) values for control versus OA ACLs based on model fitting results through contrast fitting approach, evaluating the significance of estimated logFC values by Wald test; adjusting the effects of multiple tests using false discovery rate (FDR) [32] approach to obtain FDR adjusted P-values.

### Pathway analysis of differentially expressed miRNAs and their predicted targets

Potential biological associations of the DE miRNAs in OA ACL were identified using Ingenuity Pathway Analysis (IPA) (IPA, Qiagen Redwood City, USA) ‘Core Analysis’. Additionally in order to identify putative miRNA targets, bioinformatic analysis was performed by uploading DE miRNA data into the MicroRNA Target Filter module within IPA. This identifies experimentally validated miRNA-mRNA interactions from TarBase, miRecords, and the peer-reviewed biomedical literature, as well as predicted miRNA-mRNA interactions from TargetScan. We applied a conservative filter at this point, using only experimentally validated and highly conserved predicted mRNA targets for each miRNA. Targets were also filtered on the cells fibroblasts and mesenchymal stem cells (as these were closest to potential cell types within ligament). ‘Core Analysis’ was then performed in IPA on the filtered mRNA target genes and their associated miRNAs. For each core analysis canonical pathways, novel networks, diseases and functions, and common upstream regulators were queried.

Additionally TOPP Gene [33] was used for overrepresentation analysis of the mRNA targets from Target Filter using Fisher’s Exact test with FDR correction. This tests whether the input mRNAs associate significantly with specific pathways and generates a list of biological process gene ontology (GO) terms. Terms with FDR adjusted P < 0.05 were summarised using REViGO [34] with allowed similarity of 0.4 and visualised using Cytoscape [35].

### Statistical analysis

The heatmap, volcano and principle component analysis (PCA) plots were made using MetaboAnalyst 3.5 (http://www.metaboanalyst.ca) which uses the R package of statistical computing software.30 [36].

## RESULTS

### Sample assessment

The ages of the control group (age, mean ± standard deviation (48 ± 2.16)) and ACLs derived from OA joints (74.7 ± 5.42) were significantly different (p<0.05) (Supplementary Figure 1). Summary of all donors’ information is provided in Supplementary Table 1.

### Analysis of small RNA sequencing data

We identified a total 590 miRNAs, 226 snoRNAs and 100 small nuclear (snRNAs) in the samples (with greater than 10 counts per million (CPM) in each samples). There were 184 differentially expressed sncRNAs identified (FDR<0.05) and at least 10 CPM in each sample. The categories of RNA identified are in Figure 1A and included miRNAs, snoRNAs and small nuclear RNAs (snRNAs). PCA revealed that the ACLs derived from non-OA joints (control) were clustered together and could be clearly separated from the ACLs derived from OA knee joints (Figure 1B).

**Figure 1.**
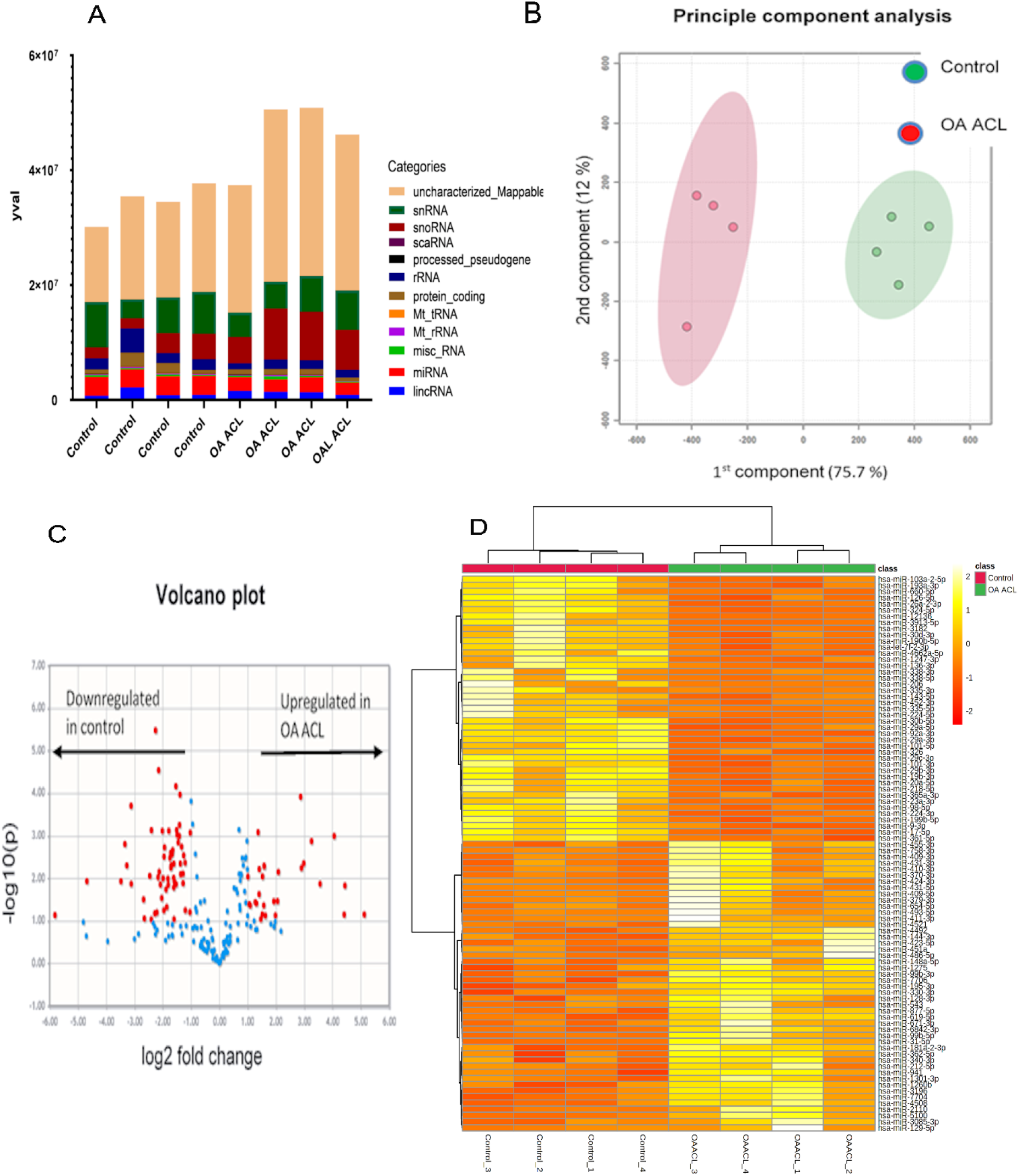
Overview of HiSeq transcriptomics data between control and diseased osteoarthritic (OA) human anterior cruciate ligament (ACL). A. Categories of RNAs identified in control and diseased OA ACL B) Principle component analysis revealed that small non-coding RNAs (sncRNA) between control and diseased ACL were distinctly grouped. C) Volcano plot demonstrate significant (FDR < 0.05) differentially expressed sncRNAs (red dots) with a fold-change of 1.4. D. Heatmap representation of the small non-coding RNA reads from control to OA ACL. Columns refer to the control and OA ACL samples and rows of miRNAs identified with their Ensembl identification. Heatmap was generated using log-transformed normalised read counts, normalisation was performed by EdgeR’s trimmed mean of M values. The colour of each entry is determined by the number of reads, ranging from yellow (positive values) to red (negative values).

Of the 184 snRNAs there were 68 DE snoRNAs (64 reduced in OA and 4 increased in OA), 26 DE snRNAs (24 reduced in OA and 2 increased in OA) and 90 DE miRNAs (43 reduced in OA and 47 increased in OA) (FDR<0.05 and greater than 10 CPM in all samples) (Figure 1C, Supplementary Table 2). The most DE miRNAs are in Table 1, with snRNA and snoRNAs in Table 2. We further generated a heatmap of the DE sncRNAs for miRNAs (Figure 1D) and snRNAs and snoRNAs (Supplementary Figure 2).

**Figure 2.**
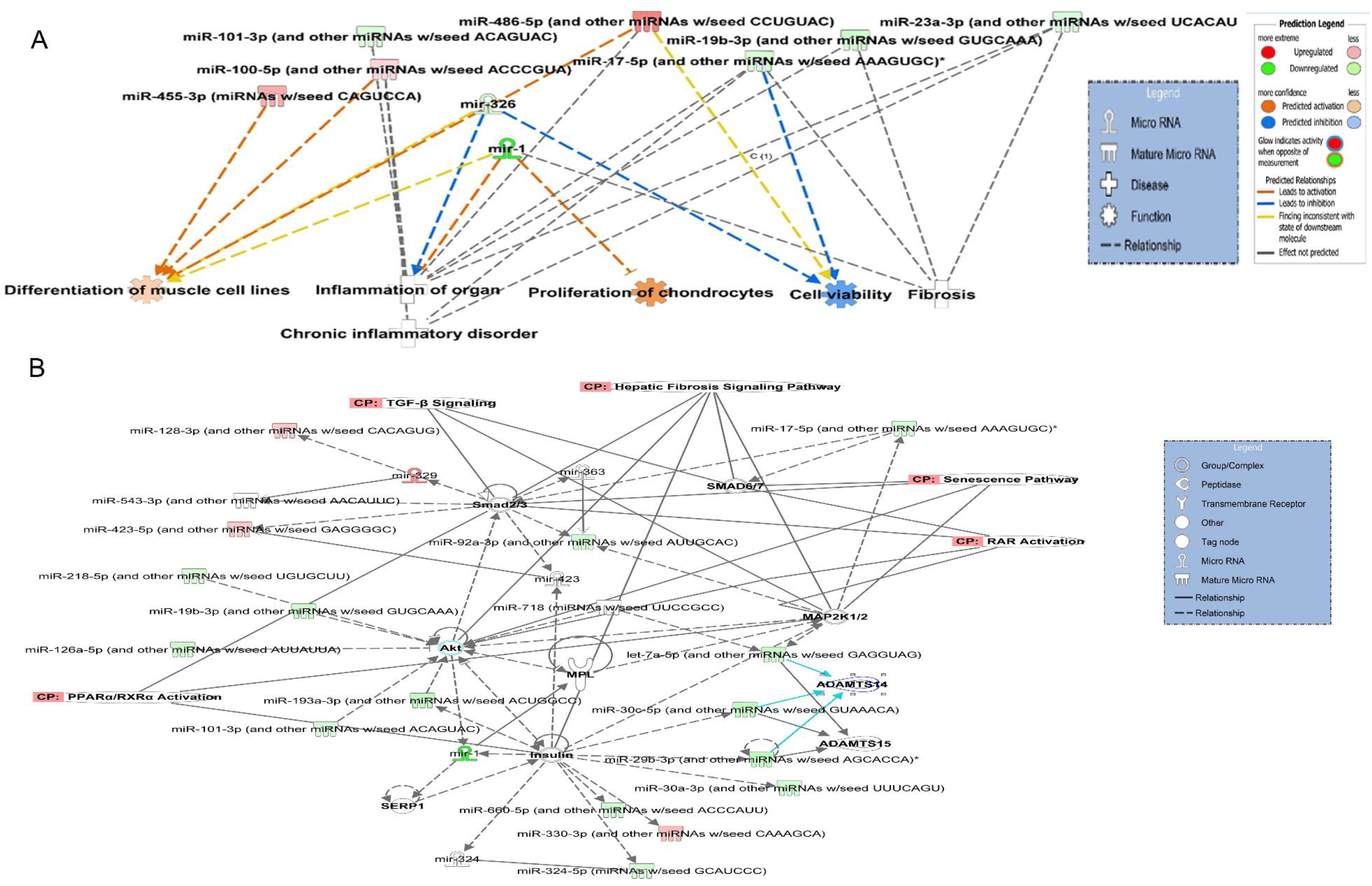
Ingenuity Pathway Analysis (IPA) derived functions of differentially expressed microRNAs in diseased osteoarthritic (OA) anterior cruciate ligament (ACL). A. IPA identified that cellular functions differentiation of muscle, inflammation, proliferation, cell viability and fibrosis were associated with the differentially expressed microRNAs. Figures are graphical representations between molecules identified in our data in their respective networks. Red nodes, upregulated in OA, and green nodes, downregulated gene expression in OA. Intensity of colour is related to higher fold-change. Legends to the main features in the networks are shown. Functions colour is dependent on whether it is predicted to be activated or inhibited. B. Top network identified with canonical pathways overlaid for fibrosis, senescence, TGFβ signalling, RAR activation and PPAR/RXR activation.

**Table 1.**
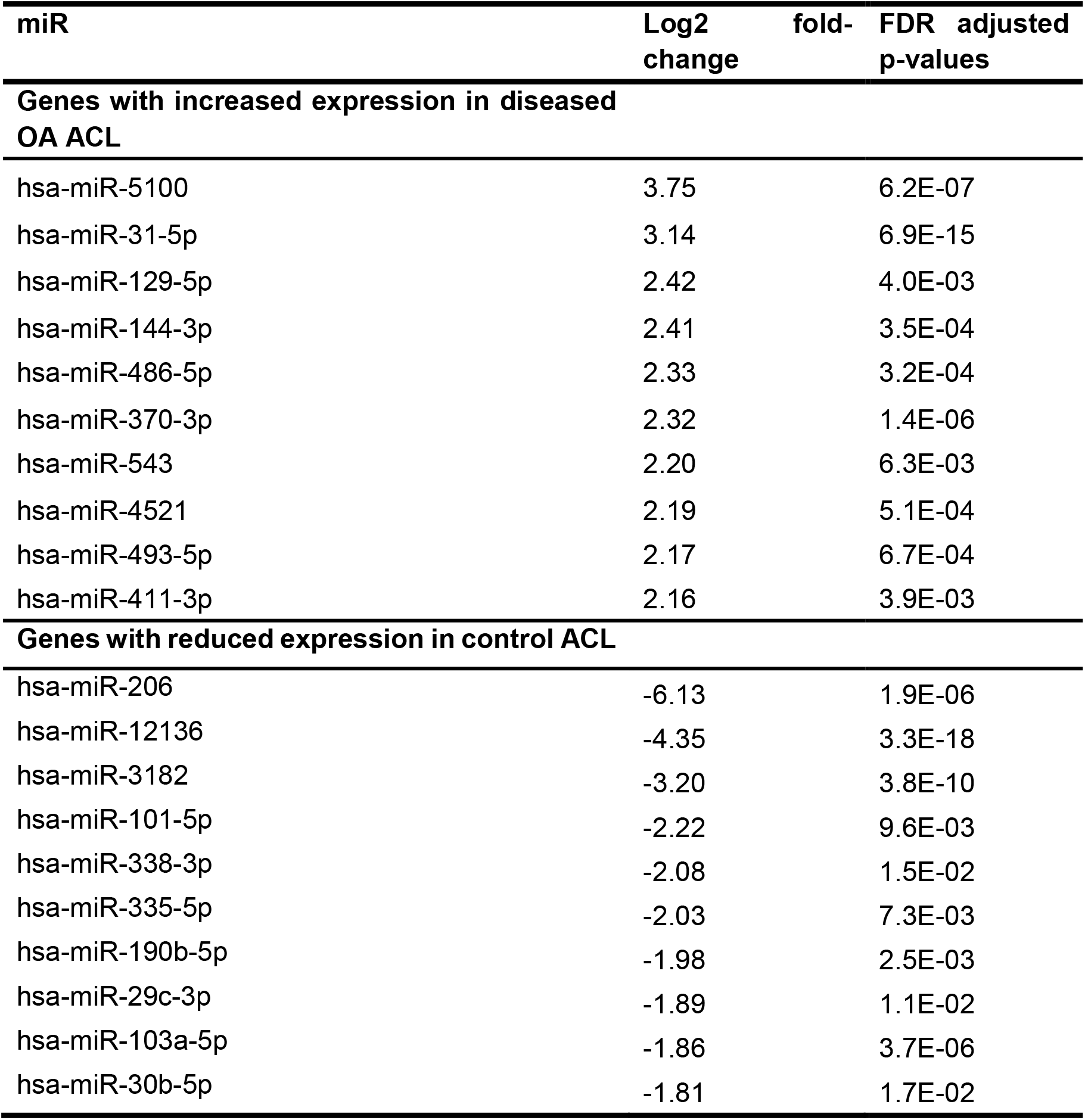
Differentially expressed miRs with the highest and lowest log2 fold-change when comparing from control versus diseased OA anterior cruciate ligament (ACL).

**Table 2.** Small-non coding RNAs (small nucleolar RNAs (snoRNAs) and small nuclear RNA (sncRNA) identified as differentially expressed between control and anterior cruciate ligaments (ACLs) derived from osteoarthritic joints.

| Name | Family | Action | Log2 fold-change | FDR adjusted p-value | Higher |
| --- | --- | --- | --- | --- | --- |
| SNORD114 | C/D BOX | Site-specific 2'-O-methylation | 3.60 | 4.7E-07 | OA<br>ACL |
| SNORD113 | C/D BOX | Site-specific 2'-O-methylation | 2.85 | 9.8E-05 | OA<br>ACL |
| RNU6 | Spliceosome | Complex of snRNA and protein subunits that removes introns from a transcribed pre-mRNA | 2.85 | 4.8E-03 | OA<br>ACL |
| SNORD72 | C/D BOX | Site-specific 2'-O-methylation | 1.83 | 4.2E-02 | OA<br>ACL |
| RNVU1-19 | Spliceosome | Complex of snRNA and protein subunits that removes introns from a transcribed pre-mRNA | 1.58 | 4.8E-02 | OA<br>ACL |
| RNU7-19P | Spliceosome | Complex of snRNA and protein subunits that removes introns from a transcribed pre-mRNA | -7.61 | 4.0E-07 | Control<br>ACL |
| RNU4-59P | Spliceosome | Complex of snRNA and protein subunits that removes introns from a transcribed pre-mRNA | -4.90 | 1.4E-33 | Control<br>ACL |
| SNORA36B | H/ACA box | H/ACA family of pseudouridylation guide snoRNAs | -4.25 | 2.7E-06 | Control<br>ACL |
| SNORA53 | H/ACA box | H/ACA family of pseudouridylation guide snoRNAs | -3.68 | 5.1E-15 | Control<br>ACL |
| SNORA73B | H/ACA box | H/ACA family of pseudouridylation guide snoRNAs | -3.61 | 3.9E-05 | Control<br>ACL |

### Pathway analysis of differentially expressed miRNAs

To explore potential biological associations of the 90 DE miRNAs in ACLs derived from OA knee joints we undertook an IPA ‘Core Analysis’. Network-eligible molecules were overlaid onto molecular networks based on information from Ingenuity Pathway Knowledge Database. Networks were then generated based on connectivity. Interesting features were determined from the gene networks inferred. Significant cellular functions deduced by the DE miRNAs included differentiation of muscle(P<0.001), inflammation (P<1.42E-10), proliferation of chondrocytes (P<0.03), fibrosis (P<0.001) and cell viability (P<0.03) (Figure 2A). The top scoring network identified was ‘Organismal Injury and Abnormalities’ (score 43) and included OA-related miRNAs such as miR-206, miR-101, let-7f, miR-455, miR-29b and miR-29c (Figure 2B).

### Pathway analysis on target mRNA genes of the differentially expressed miRNAs

Next, we undertook analysis to determine the mRNA targets of the DE miRNAs. 90 miRNAs that were DE in ACLs derived from OA knee joints compared to controls were initially input into MicroRNA Target Filter. Once a conservative filter was applied (only miRNAs with experimental or highly predicted targets), 529 mRNAs were putative targets (Supplementary Table 3). These mRNAs were then input into IPA core analysis and all results summarised in Supplementary Table 3. The top canonical pathways for target mRNAs of DE miRNAs in OA ACL are in Table 3. Two of the most significant of which were the osteoarthritis pathway (P<2.3E-23) and hepatic fibrosis (P<3.1E-32) (Figure 3). The most significant upstream regulators of these mRNAs included tumour necrosis factor (P<1.3E-101) and transforming growth factor β (TGFβ) (P<8.5E-83) (Table 4). Upstream regulators represent molecules that may be responsible for the putative mRNAs in our dataset and cover the gamut of molecule types found in the literature, from transcription factors, to cytokines, chemicals and drugs. The most significant diseases and biological functions identified are shown in Table 5. The top networks identified are in Supplementary Table 3. The network ‘cellular development, movement and genes expression’ (score 41) (Figure 4A) was overlaid with significant biological processes including apoptosis (P<1.7E-85), fibrosis (P<1.2E-79), inflammation (P<3.4E-88), and necrosis (P<7.2E-88). The network ‘inflammatory disease’ (score 35) (Figure 4B) shows pertinent significant biological processes including organisation of collagen fibrils (P<3.7E-07), fibrosis (P<2.6E-14), rheumatoid arthritis (P<3.6E-06), angiogenesis (P<8.9E-09), differentiation of bone (P<5E-06), inflammation of the joint (P<8.8E-07) and cartilage development (1.5E-07).

**Table 3.**
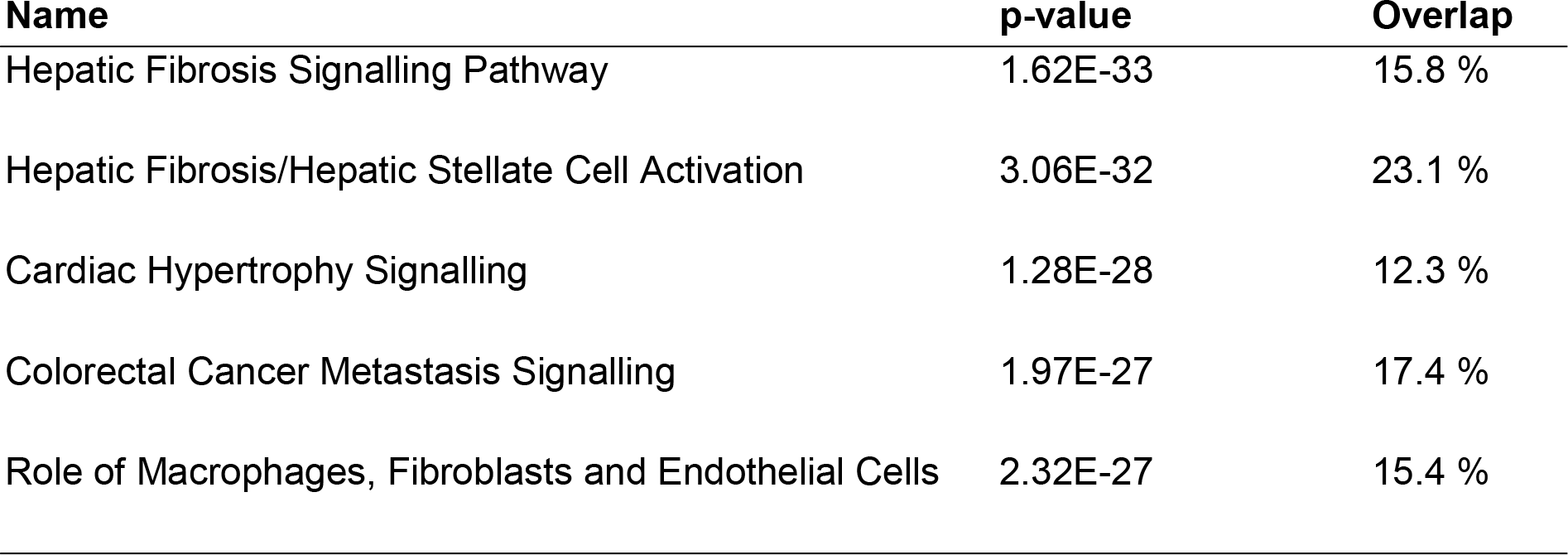
Top canonical pathways for target mRNAs of differentially expressed miRNAs in diseased OA anterior cruciate ligaments (ACLs).

| **Name** | **p-value** | **Overlap** |
| --- | --- | --- |
| Hepatic Fibrosis Signalling Pathway | 1.62E-33 | 15.8 % |
| Hepatic Fibrosis/Hepatic Stellate Cell Activation | 3.06E-32 | 23.1 % |
| Cardiac Hypertrophy Signalling | 1.28E-28 | 12.3 % |
| Colorectal Cancer Metastasis Signalling | 1.97E-27 | 17.4 % |
| Role of Macrophages, Fibroblasts and Endothelial Cells | 2.32E-27 | 15.4 % |

**Table 4.**
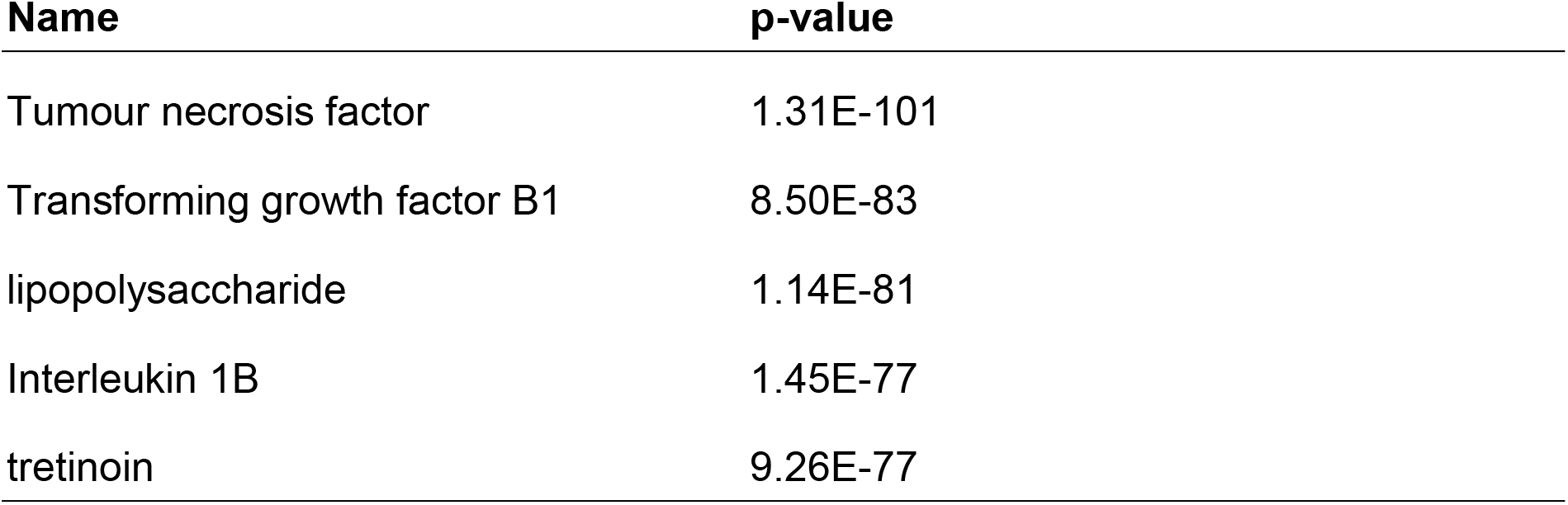
Top upstream regulators of differentially expressed miRNAs in diseased OA anterior cruciate ligaments (ACLs).

| Name | p-value |
| --- | --- |
| Tumour necrosis factor | 1.31E-101 |
| Transforming growth factor B1 | 8.50E-83 |
| lipopolysaccharide | 1.14E-81 |
| Interleukin 1B | 1.45E-77 |
| tretinoin | 9.26E-77 |

**Figure 3.**
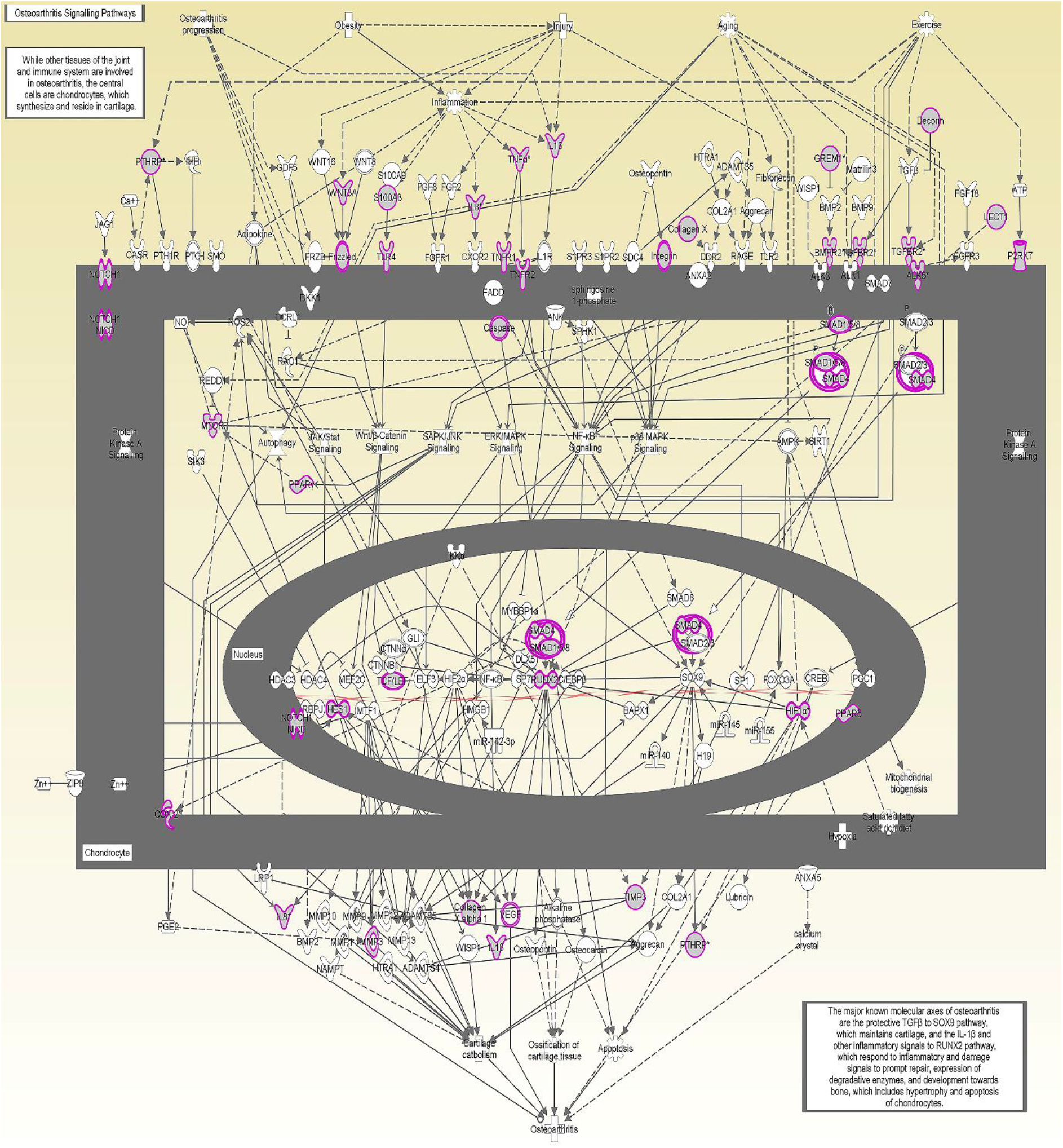
Osteoarthritis pathway targeted by predicted mRNA. The canonical pathway for osteoarthritis signalling was highly ranked (p = 2.33 E^−23^) using target mRNAs identified in Ingenuity Pathway Analysis (IPA) Targetscan from the differentially expressed miRNAs in diseased anterior cruciate ligaments (ACLs) derived from OA patients. The pathway was generated using IPA.

**Figure 4.**
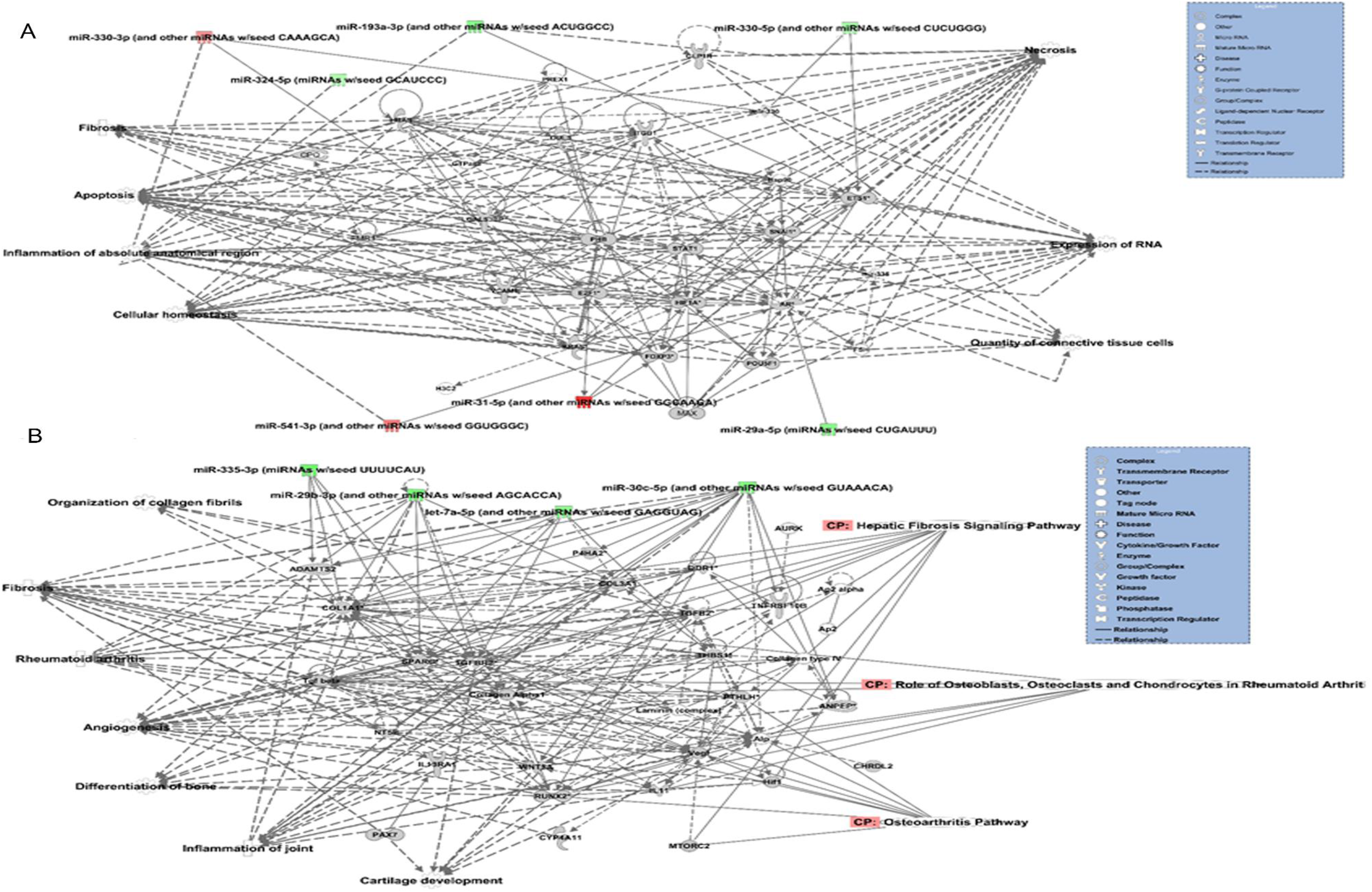
Top-scoring networks derived from the 529 putative mRNAs differentially expressed in anterior cruciate ligaments (ACLs) derived from osteoarthritic (OA) joints. Ingenuity pathway analysis (IPA) identified A. ‘Cellular development, movement and genes expression’ with a scores of 41. (B) ‘Inflammatory disease, organismal injuries and abnormalities’ with a score of 35 and within this network are molecules linked to their respective canonical pathways. Both are overlaid with pertinent significant biological functions contained in the gene sets. Figures are graphical representations between molecules identified in our data and predicted mRNA targets in their respective networks. Green nodes, downregulated in ACLs from OA joints; red nodes, upregulated gene expression in ACLs from OA joints. Intensity of colour is related to higher fold-change. Key to the main features in the networks is shown.

To obtain an overview of pathways that the putative target mRNAs were involved in the mRNAs derived from IPA were also input into the gene ontology (GO) tool TOPP Gene and the biological processes summarised in REViGO and visualised using Cytoscape (Figure 5 and Supplementary Table 4).

**Figure 5.**
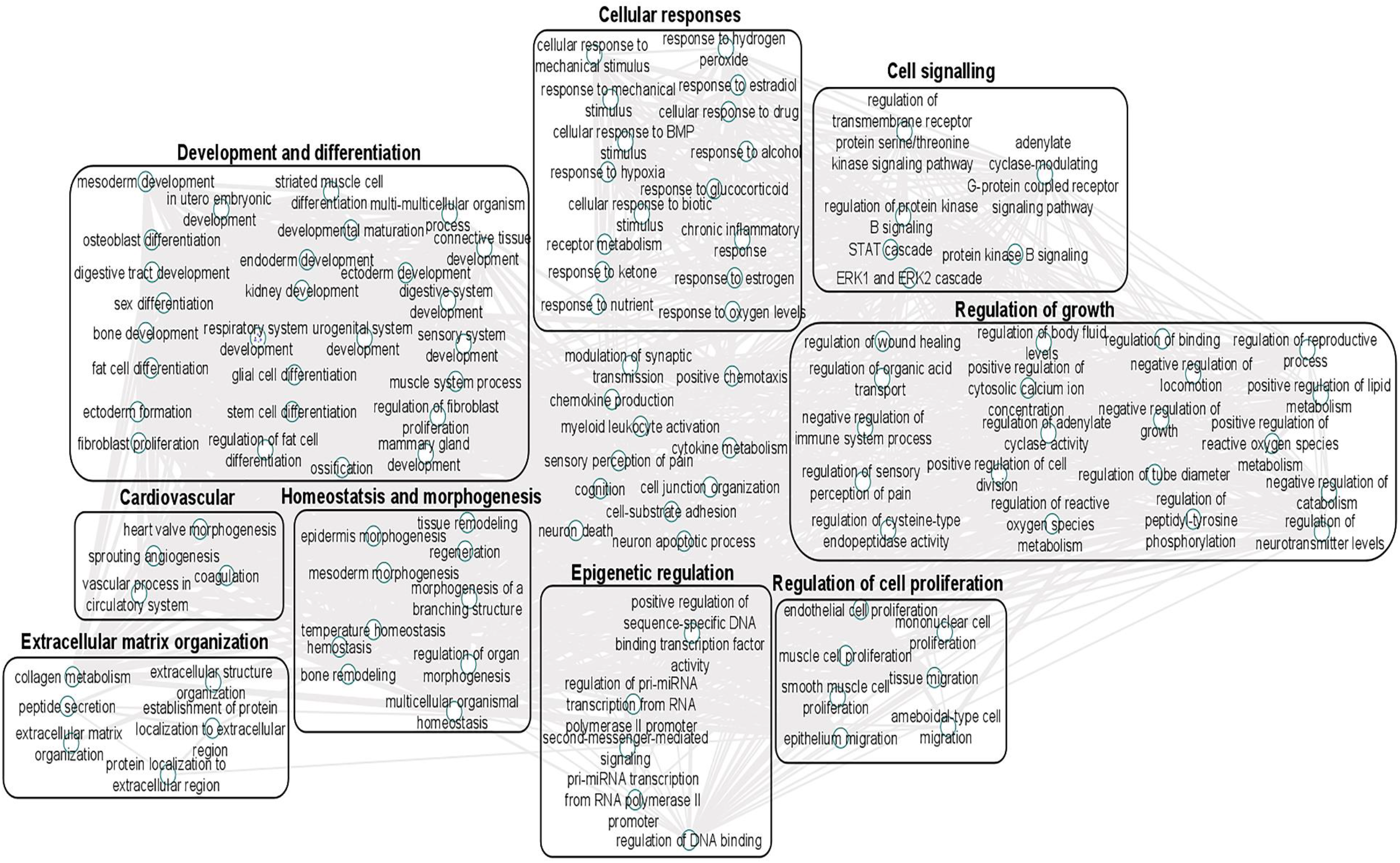
Gene ontology (GO) biological processes associated with dysregulated microRNAs targets were identified following TargetScan filter module using IPA. GO terms for biological process (FDR < 0.05) were summarized with ToppGene and visualised using REViGO and Cytoscape. Boxes represent the main clusters of biological processes that were significantly influenced by dysregulated micrRNAs between control and diseased osteoarthritic (OA) anterior cruciate ligaments (ACLs).

## DISCUSSION

The global prevalence of knee OA is currently 5% and is projected to rise with an increase in the ageing population [37]. Reports propose that there is an association between ACL degeneration and subsequent knee OA, suggesting the importance of ACL degradation and regeneration molecular mechanisms in OA pathogenesis [38]. There is also a strong correlation between ACL degeneration and subsequent knee OA [12]. One potential mechanism capable of regulating global alterations to a particular tissue is modification of sncRNA expression. To begin to elucidate the role that they play in the global changes observed in the ACL during OA and understand further the potential role of the ACL in arthritis, we undertook a non-biased approach small RNA sequencing of ACLs from OA knee joints and compared these to our control samples derived from non-OA knee joints. Whilst a study previously demonstrated DE miRNAs in ACL [20], this is the first time that, to our knowledge, small RNA sequencing has been used to interrogate both snoRNAs and miRNAs in an unbiased manner and we identified unique OA dependant signatures.

There were 68 snoRNAs, 26 snRNAs and 90 miRNAs significantly different in ACLs derived from OA knee joints and the OA status of the donor accounted for the principal variability in the data. Additional bioinformatics was performed, to analyse the biological processes and pathways affected by the differentially expressed miRNAs and in addition, the putative mRNA targets of the differentially expressed miRNAs, enhancing our understanding of the roles of the dysregulated miRNAs in OA pathogenesis.

Several of DE miRNAs found in this study, including miR-29b, miR –335, let-7f, miR-424, and miR-941 correlate to previously altered miRNAs in a study comparing ruptured ACLs to diseased OA ACLs [20]. These miRNAs were found to be correlated with cartilage development and remodelling, extracellular matrix homeostasis and inflammatory response [20]. We have found other miRNAs associated with OA including miR-206, miR-101, miR365, miR-29b and miR-29c, whose expression altered in ACLs derived from OA joints [39–41]. Pathways identified by the DE miRNAs with known functions in OA in other tissues included inflammation [42], proliferation of chondrocytes [43], and fibrosis [44]. Canonical pathways identified have roles in OA pathogenesis including senescence [45], fibrosis, TGFβ signalling [46], retinoic acid binding protein (RAR) activation [47] and peroxisome proliferator-activated receptor/retinoid X receptor (PPAR/RXR) activation [48, 49].

To address the roles of miRNAs in diseased OA ACLs, their mRNA target genes should also be taken into consideration. Therefore, in order to determine potential mRNA targets of the DE miRNAs we used Target Filter in IPA to identify predicted mRNA targets. We used conservative filters for the mRNAs identified by only choosing highly conserved predicted mRNA targets for each miRNA and by choosing relevant cell types to ACL tissue. We then undertook GO analysis and visualisation of the significant biological processes effected with online tools. Using ToppGene, Revigo and Cytoscape for visualisation, we showed an overview of the essential biological processes these mRNA target genes were involved in, including extracellular matrix organisation, epigenetic regulation, cell signalling, cell growth and proliferation. In IPA, additional functions affected by these genes, known to have a role in OA pathogenesis and therefore with a potential role in OA ACLs were highlighted including apoptosis [50], fibrosis [44], inflammation of the joint [42], necrosis [51], organisation of collagen fibrils [12], angiogenesis [52], differentiation of bone [53] and cartilage development [54]. Canonical pathway analysis was also performed. We found that many pathways enriched by the putative target genes were essential for OA pathogenesis, including the ‘osteoarthritis pathway’. The significant mRNA targets in this pathway included those involved in inflammation (TNF, IL1, IL8), Wnt signalling (WNt3A, Frizzled, TCF/LEF), TGFβ and SMAD signalling (TGFβR2, SMAD1, –4, –5, –8), hypoxia (HIF1α), and mammalian target of rapamycin signalling (MTOR). Additionally downstream targets of these signalling pathways with known roles in OA pathogenesis were identified and included matrix metalloproteinase-3 [55], tissue inhibitor of metalloproteinase-3 [56], and collagen X α1 [57]. These findings indicate the potential importance of these pathways in ACL degeneration associated with OA. Hepatic fibrosis was the most significant canonical pathway identified from the putative mRNAS together with the DE miRNAs in our study. Synovial fibrosis is often found in OA [44] and fibrosis has previously been described in OA joints following ACL injury [58]. Furthermore TGFβ, one of the most significant upstream regulator in our mRNA target gene analysis, is the master regulator of fibrosis [57]. Many TGFβ-related genes including TGFβ2, TGFβ3, TGFβR1, TGFβR2 and TGFβR3 were predicted targets of the DE miRNAs including miR-98–5p, miR-101–3p, miR-128–5p, miR-136–3p, miR-17-5p; strongly implicating it in the fibrosis evident in the diseased ACLs in OA.

Another class of snRNAs, snoRNAs, were altered in the OA ACLs in our study. This conserved class of non-coding RNAs are principally characterised as guiding site-specific post-transcriptional modifications in ribosomal RNA [59]. Furthermore snoRNAs can modify and/or interact with additional classes of RNAs including other snoRNAs, transfer RNAs and mRNAs [60]. A reliable modification site has been assigned to 83% of the canonical snoRNAs, with 76 snoRNAs described as orphan, meaning they act in an unknown or unique manner [61]. Novel functions reported for snoRNAs including the modulation of alternative splicing [62], involvement in stress response pathways, [63] and the modulation of mRNA 3′end processing [64]. Like miRNAs, snoRNAs are emerging as important regulators of cellular function and disease development [65], in part due to their ability to fine-tune the ribosome to accommodate changing requirements for protein production during development, normal function and disease [66].

We have previously identified a role for snoRNAs in cartilage ageing and OA [24] and their potential use as biomarkers for OA [27]. Furthermore, others identified that the snoRNAs, SNORD38 and SNORD48, are significantly elevated in the serum of patients developing cartilage damage a year following ACL injury and serum levels of SNORD38 were greatly elevated in patients who develop cartilage damage after ACL injury suggesting SNORD38 as a serum biomarker for early cartilage damage [67]

Interestingly, we found an increase in SNORD113 and SNORD114 in diseased OA ACLs. These snoRNAs are located in imprinted human loci and may play a role in the evolution and/or mechanism of epigenetic imprinting [61].They belong to the C/D box class of snoRNAs and most of the members of the box C/D family direct site-specific 2’-O-methylation of substrate RNAs. However, SNORD113 and SNORD114 differ from C/D box snoRNAs in their tissue specific expression profiles (including in fibroblasts, osteoblasts and chondrocytes [61] and the lack of complementarity to any RNA. As a result, they are not predicted to guide to 2’O-methylation but have novel, unknown roles [61]. Additionally SNORD113–1 functions as a tumour suppressor in hepatic cell carcinoma by reducing cell growth and inactivating the phosphorylation of ERK1/2 and SMAD2/3 in MAPK/ERK and TGF-β pathways [68]. We have previously identified that SNORD113–1 is also increased in OA human knee cartilage but reduced in ageing human knee cartilage, whilst SNORD114 increases in OA knee cartilage [24].

SNORD72 was increased in diseased OA ACLs. In hepatocellular carcinoma, the overexpression of SNORD72 was found to enhance cell proliferation, colony formation and invasion by stabilising Inhibitor of differentiation (ID) genes which are a basic helix-loop-helix (bHLH) transcription factors [69]. The ID family genes have been shown to play a role in cell proliferation and angiogenesis [70]. The lack of a DNA binding domain, results in inhibition of the binding of other transcription factors to DNA in a dominant negative fashion [71]. The expression of some members of this family in rheumatoid arthritis synovium suggests they may have a role in human inflammatory disease [72]. Whilst the downstream signalling of snoRNAs is principally unknown, snoRNAs regulate ribosome biogenesis [73]. However a subclass of orphans do not have complimentary RNA sequences [74]. Mao Chet et al., found that ribosome biogenesis was not affected following SNORD72 overexpression implying it exerts functionality in other ways [69]. Therefore whilst some snoRNAs can regulate the expression of RNAs [75], others can reduce the gene stability [74] or directly activate or suppress enzymes [76]. Together our snoRNA findings indicate that changes in ACL snoRNA expression could have important implications in knee OA through both canonical and non-canonical roles.

Our study has a number of limitations due to availability of human ACL tissue. There was also a mild imbalance between the sexes in the two groups with most of the OA derived ACLs coming from males but all the control group being sourced from females. In human tendon, we have previously demonstrated that males and females are transcriptionally different and gene expression in aged cells moves in opposite directions [26]. Ligament degeneration has also been demonstrated to be influenced by lower concentrations of sex hormones in young female athletes [77]. Macroscopic grading of tissues were not performed due to limited images of diseased OA ACL samples, however images of control samples demonstrated healthy knee joint cartilage with no signs of ACL degeneration. Finally, there were age discrepancies between the two groups and so we cannot discount an age effect on sncRNAs expression.

In conclusion, our study revealed alterations in a number of classes of snRNAs in ACL tissues derived from patients with knee OA compared to apparently healthy ACLs from non-OA joints. Our functional bioinformatic analyses suggests that the dysregulated miRNAs may regulate cartilage development and remodelling, collagen biosynthesis and degradation, ECM homeostasis and pathology by interacting with their targets. Uniquely we also demonstrate that snoRNAs may also have a role in ACL degeneration. Collectively, our study provides novel insight into the ACL related sncRNA dysregulation in patients with OA.

## Data Availability

The datasets supporting the conclusions of this article are included within the article and its additional files.

## Acknowledgement

Mandy Peffers funded through a Wellcome Trust Intermediate Clinical Fellowship (107471/Z/15/Z). This work was also supported by Institute of Ageing and Chronic disease (Comerford) and by the MRC and Versus Arthritis as part of the Medical Research Council Versus Arthritis Centre for Integrated Research into Musculoskeletal Ageing (CIMA) [MR/R502182/1]. The MRC Versus Arthritis Centre for Integrated Research into Musculoskeletal Ageing is a collaboration between the Universities of Liverpool, Sheffield and Newcastle. The authors would like to acknowledge, Dr Pieter Emans, Maastricht University Orthopeadic surgeon for collating OA diseased samples.

## Authors’ contributions

MP, EC and YAK designed and coordinated the study. TW collected the samples. YF processed the samples for small RNA sequencing. MP and YA conducted the statistical analysis and drafted the manuscript. All authors revised the draft critically read and approved the final submitted manuscript.

## Supplementary Figures and Tables

**Supplementary Figure 1.** Age groups between control anterior cruciate ligament (ACL) samples and diseased OA ACL samples.

**Supplementary Figure 2.** Heatmap representation of the differentially expressed snoRNAs and sncRNAs small non-coding RNA reads from control to OA anterior cruciate ligament (ACL)

**Supplementary Table 1.** Donors’ age, gender and ethnicity information.

**Supplementary Table 2.** Differentially expressed sncRNAs between control and OA anterior cruciate ligament (ACL) samples with FDR<0.05 and reads greater than 10 counts per million (CPM).

**Supplementary Table 3.** Ingenuity Pathway Analysis (IPA) of differentially expressed microRNAs target genes between control and OA anterior cruciate ligament (ACL) and IPA of mRNA core analysis of putative mRNAs targets

**Supplementary Table 4.** Gene ontology (GO) of the top biological processes gene ontology of putative mRNA targets between control and OA anterior cruciate ligament (ACL) summarised in REViGO tool.

